# Assessing GPT-4 Multimodal Performance in Radiological Image Analysis

**DOI:** 10.1101/2023.11.15.23298583

**Authors:** Dana Brin, Vera Sorin, Yiftach Barash, Eli Konen, Girish Nadkarni, Benjamin S Glicksberg, Eyal Klang

## Abstract

**Objectives:** This study aims to assess the performance of OpenAI’s multimodal GPT-4, which can analyze both images and textual data (GPT-4V), in interpreting radiological images. It focuses on a range of modalities, anatomical regions, and pathologies to explore the potential of zero-shot generative-AI in enhancing diagnostic processes in radiology.

**Methods:** We analyzed 230 anonymized emergency room diagnostic images, consecutively collected over one week, using GPT-4V. Modalities included ultrasound (US), computerized tomography (CT) and X-ray images. The interpretations provided by GPT-4V were then compared with those of senior radiologists. This comparison aimed to evaluate the accuracy of GPT-4V in recognizing the imaging modality, anatomical region, and pathology present in the images.

**Results:** GPT-4V identified the imaging modality correctly in 100% of cases (221/221), the anatomical region in 87.1% (189/217), and the pathology in 35.2% (76/216).

However, the model’s performance varied significantly across different modalities, with anatomical region identification accuracy ranging from 60.9% (39/64) in US images to 97% (98/101) and 100% (52/52) in CT and X-ray images (p<0.001).

Similarly, Pathology identification ranged from 9.1% (6/66) in US images to 36.4% (36/99) in CT and 66.7% (34/51) for X-ray images (p <0.001).

These variations indicate inconsistencies in GPT-4V’s ability to interpret radiological images accurately.

**Conclusion:** While the integration of AI in radiology, exemplified by multimodal GPT-4, offers promising avenues for diagnostic enhancement, the current capabilities of GPT-4V are not yet reliable for interpreting radiological images. This study underscores the necessity for ongoing development to achieve dependable performance in radiology diagnostics.

## Introduction

Artificial Intelligence (AI) is transforming medicine, offering significant advancements especially in data-centric fields like radiology. Its ability to refine diagnostic processes and improve patient outcomes marks a revolutionary shift in medical workflows.

Radiology, heavily reliant on visual data, is a prime field for AI integration(1). AI’s ability to analyze complex images offers significant diagnostic support, potentially easing radiologist workloads by automating routine tasks and efficiently identifying key pathologies(2). The increasing use of publicly available AI tools in clinical radiology has integrated these technologies into the operational core of radiology departments(3–5).

Among AI’s diverse applications, Large Language Models (LLMs) have gained prominence, particularly GPT-4 from OpenAI, noted for its advanced language understanding and generation(6–15). A notable recent advancement of GPT-4 is its multimodal ability to analyze images alongside textual data (GPT-4V)(16). The potential applications of this feature can be substantial, specifically in radiology where the integration of imaging findings and clinical textual data is key to accurate diagnosis. Thus, the purpose of this study was to evaluate the performance of GPT-4V for the analysis of radiological images across various imaging modalities and pathologies.

## Methods

A Sheba Medical Center Institutional Review Board (IRB) approval was granted for this study. The IRB committee waived informed consent.

### Dataset Selection

In this retrospective study, we conducted a systematic review of all imaging examinations recorded in our hospital’s Radiology Information System (RIS) during the first week of October 2023. The study specifically focused on cases presenting to the emergency room (ER).

Our inclusion criteria included complexity level, diagnostic clarity and case source. Regarding level of complexity, we selected ‘resident-level’ cases, defined as those that are typically diagnosed by a first-year radiology resident. These are cases where the expected radiological signs are direct and the diagnoses are unambiguous. Regarding diagnostic clarity, we included ‘clear-cut’ cases with a definitive radiologic sign and diagnosis stated in the original radiology report, which had been made with a high degree of confidence by the attending radiologist. These cases included pathologies with characteristic imaging features that are well-documented and widely recognized in clinical practice. Only selected cases originating from the ER were considered, as these typically provide a wide range of pathologies, and the urgent nature of the setting often requires prompt and clear diagnostic decisions.

We deliberately excluded any cases where the radiology report indicated uncertainty. This ensured the exclusion of ambiguous or borderline findings, which could introduce confounding variables into the evaluation of the AI’s interpretive capabilities. The aim was to curate a dataset that would allow for a focused assessment of the AI’s performance in interpreting imaging examinations under clear, clinically relevant conditions without the potential bias of complex or uncertain cases.

An attending body imaging radiologist, together with a 2^rd^ year radiology resident, conducted the case screening process based on the predefined inclusion criteria. They consensually agreed on the selection of cases for the study.

A total of 230 images were selected, which represented a balanced cross-section of modalities including computed tomography (CT), ultrasound (US), and X-ray (**Table 1**). These images spanned various anatomical regions and pathologies, chosen to reflect a spectrum of common and critical findings appropriate for resident-level interpretation.

**Table 1:** Aggregated Data of Anatomical Regions and Pathologies by Imaging Modality.

| Modality and anatomical region | Total number of images<br>n | Normal<br>n (%) | Pathology<br>n (%) | Pathologies list |
|---|---|---|---|---|
| **All** | 230 | 111<br>(48.3) | 119<br>(51.7) |  |
| **X-ray**<br>Chest | 53 | 22<br>(41.5) | 31<br>(58.5) | Mass, cardiac pacemaker, pulmonary edema, various forms of consolidation, pleural effusion, pneumothorax, sternotomy |
| **CT** | 103 | 47<br>(45.6) | 56<br>(54.4) |  |
| Abdomen and pelvis | 47 | 21<br>(44.7) | 26<br>(55.3) | Cholelithiasis, cholecystitis, small bowel obstruction, bowel perforation (pneumoperitoneum), liver metastases, liver cysts, ascites, pyelonephritis, colonic obstruction, appendicitis, diverticulitis, colitis |
| Spine | 13 | 6<br>(46.2) | 7<br>(53.8) | Fracture, degenerative changes, sclerotic lesion |
| Head | 43 | 20<br>(46.5) | 23<br>(53.5) | Parenchymal hemorrhage, subdural hemorrhage, hydrocephalus, ventriculoperitoneal shunt, acute stroke |
| **US** | 74 | 42<br>(56.8) | 32<br>(43.2) |  |
| Kidney | 46 | 26<br>(56.5) | 20<br>(43.5) | Hydronephrosis, nephrolithiasis, renal cyst |
| RUQ | 10 | 5<br>(50) | 5<br>(50) | Cholecystitis, cholelithiasis, liver metastasis |
| Testicles | 18 | 11<br>(61.1) | 7<br>(38.9) | Testicular Mass |

\* Some cases included more than one pathology, for example a consolidation and a pleural effusion.
Abbreviations: ultrasound (US), computed tomography (CT), right upper quadrant (RUQ).

To uphold the ethical considerations and privacy concerns, each image was anonymized to maintain patient confidentiality prior to analysis. This process involved the removal of all identifying information, ensuring that the subsequent analysis focused solely on the clinical content of the images.

### AI Interpretation with GPT-4 Multimodal

Using OpenAI’s API, GPT-4V was prompted to analyze each image. We asked for identification of the modality, anatomical region, and pathology, in a JSON format, to allow efficient analysis of the results. The specific prompt used was “*We are conducting a study to evaluate GPT-4 image recognition abilities in healthcare. Identify the condition and describe key findings in the image. Please return the answer in a JSON format, and specify the modality, anatomical region, and pathology. {“modality”:<type of imaging modality>, “anatomical_region”: <anatomical region of the image>, “pathology”: <pathology in the image, or normal if none is shown>}*.*”* The attending radiologist and the resident reviewed the AI interpretations in consensus and compared them to the imaging findings.

To evaluate GPT-4V’s performance, we checked for the accurate recognition of modality type, anatomical location, and pathology identification. Errors were classified as omissions and hallucinations.

### Data Analysis

The analysis was performed using Python version 3.10. Statistical significance was determined using a p-value threshold of less than 0.05.

The primary metrics were the model accuracies of modality, anatomical region, and overall pathology diagnosis. These metrics were calculated per modality, as correct answers out of all answers provided by GPT-4V. The overall pathology diagnostic accuracy was calculated as the sum of correctly identified pathologies and the correctly identified normal cases out of all cases answered. A qualitative analysis of GPT-4V answers was also performed.

Chi-square tests were employed to assess differences in the ability of GPT-4V to identify modality, anatomical locations, and pathology diagnosis across imaging modalities.

## Results

### Distribution of Imaging Modalities

The dataset consists of 230 diagnostic images categorized by modality (CT, X-ray, US), anatomical regions and pathologies. The results are summarized in **Table 1**. Overall, 119 images (51.7%) were pathological, 111 cases (48.3%) were normal.

### Excluded Cases

During our analysis, there were instances where GPT-4V failed to provide a response related to the image modality, anatomical region, or pathology. The output in such cases was either “unable to provide diagnoses or interpret medical images” or simply “unknown”, which applied to either the entire question or to specific sections of the JSON structure (modality, anatomy, or pathology). As a result, these instances were omitted from the final analysis. Specifically, we excluded 9/230 cases for modality, 13/230 for anatomical region, and 14/230 for pathology.

### GPT-4V Performance in Imaging Modality and Anatomical Region Identification

GPT-4V provided an answer regarding the imaging modality in 221/230 cases. GPT-4V demonstrated a 100% (221/221) accuracy rate for identification of the imaging modalities, across CT, US, and X-ray images (**Table 2**).

**Table 2:** GPT-4 Modality and Anatomy Identification Accuracy. Identified/Total (%)

| **Modality** | **Modality** | **Anatomical region** |
|---|---|---|
| **CT** | 101/101 (100%) | 98/101 (97%) |
| **X-ray** | 52/52 (100%) | 52/52 (100%) |
| **US** | 68/68 (100%) | 39/64 (60.9%) |
| **Total** | 221/221 (100%) | 189/217 (87.1%) |

Abbreviations: ultrasound (US), computed tomography (CT).

When asked about the anatomical region, GPT-4V provided an answer in 217/230 cases. The model correctly identified 100% (52/52), 97% (98/101) and 60.9% (39/64) of X-ray, CT, and US images (p<.001), with an overall 87.1% (189/217) accuracy (**Table 2**).

### GPT-4V Performance in Overall Pathology Diagnostic Accuracy

GPT-4V answered 216/230 cases when asked about the presence of pathologies, out of which 111 were pathological and 105 normal cases. GPT-4V demonstrated an accuracy of 35.2% (76/216) in pathology diagnosis, which differed notably across imaging modalities (**Table 3**). Accuracy from diagnoses based on X-ray images was 66.7% (34/51), from CT images was 36.4% (36/99) and from US images was 9.1% (6/66). The difference between the modalities was statistically significant (p <0.001). Examples of cases from the GPT-4V image analysis are presented in **Figures 1-6**.

**Table 3:**
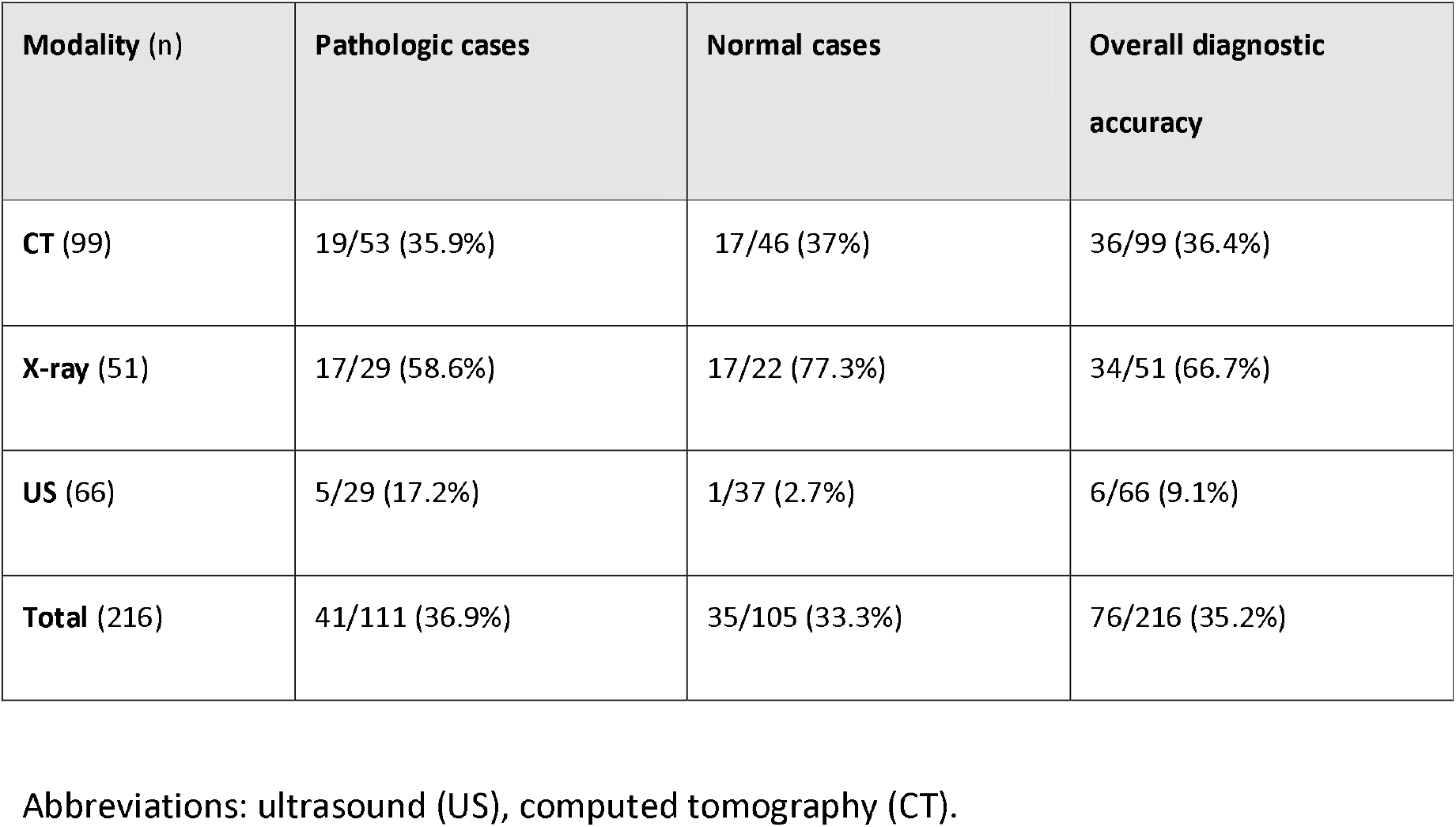
GPT-4 Pathology Identification Accuracy. Identified/Total (%)

**Figure 1.**
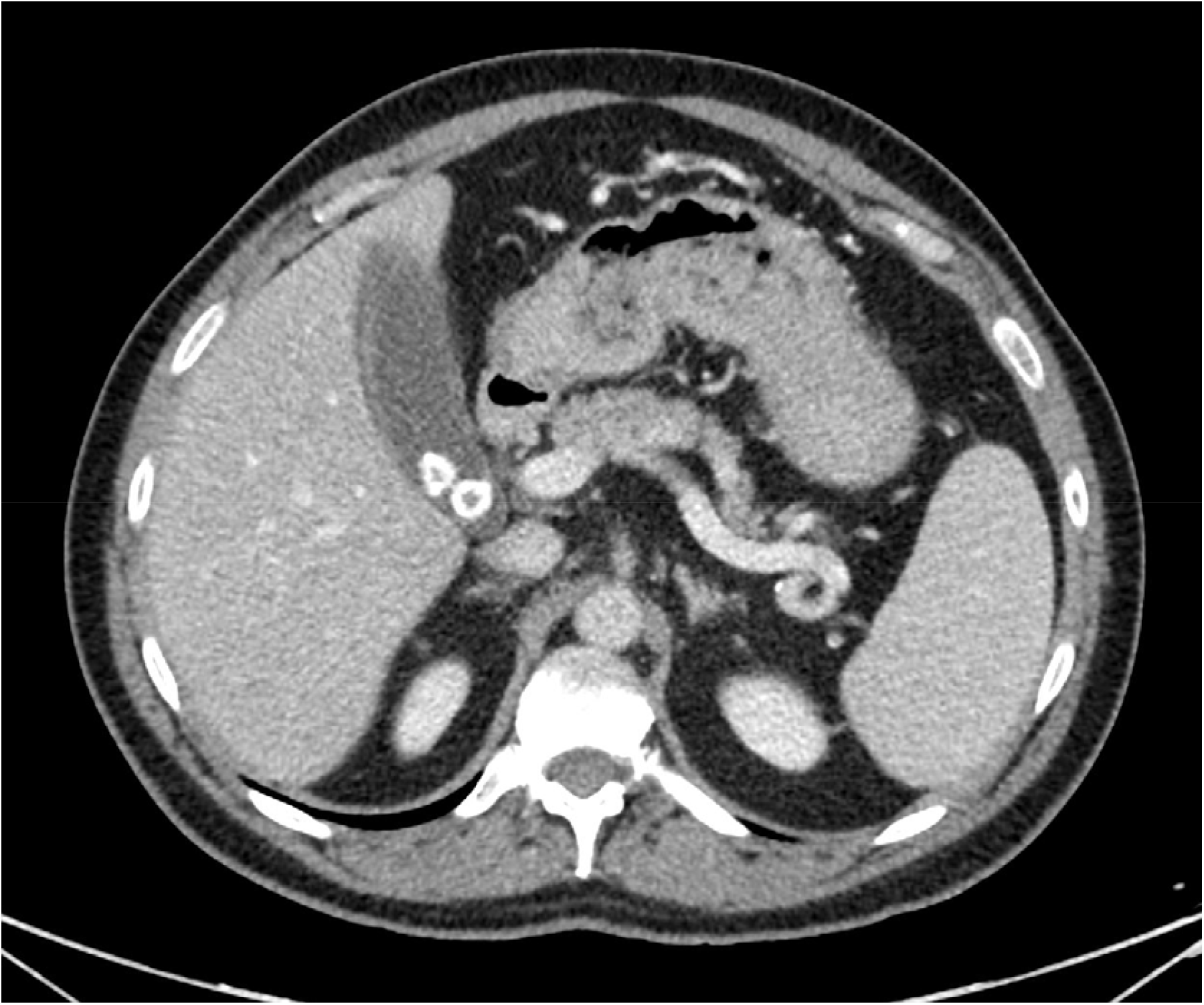
CT scan of the abdomen showing cholelithiasis. GPT-4V correctly identify the image modality (“CT scan”) and the anatomical location (“Abdomen”). The pathology identify by GPT-4V is “Splenic laceration with subcapsular hematoma and hemoperitoneum”, meaning it missed the correct pathology of cholelithiasis and hallucinated the splenic pathology. GPT-4V’s full answer: ‘‘‘json } “modality”: “CT scan,” “anatomical_region”: “Abdomen,” “pathology”: “Splenic laceration with subcapsular hematoma and hemoperitoneum” { ‘‘‘

Of the correct cases, in ten X-rays and two CT images, despite the correctly identified pathology, the description of the pathology was not accurate and contained errors related to the meaning or location of the pathological finding. An example of this is shown in **Figure 6**.

Out of the incorrect pathologic cases, 25.7% (18/70) were due to omission of the pathology and misclassifying the image as normal (**Figure 2**), and 57.1% (40/70) were due to hallucination of an incorrect pathology (**Figure 3**). The rest were due to incorrect identification of the anatomical region (17.1%, 12/70)(**Figure 5**).

**Figure 2.**
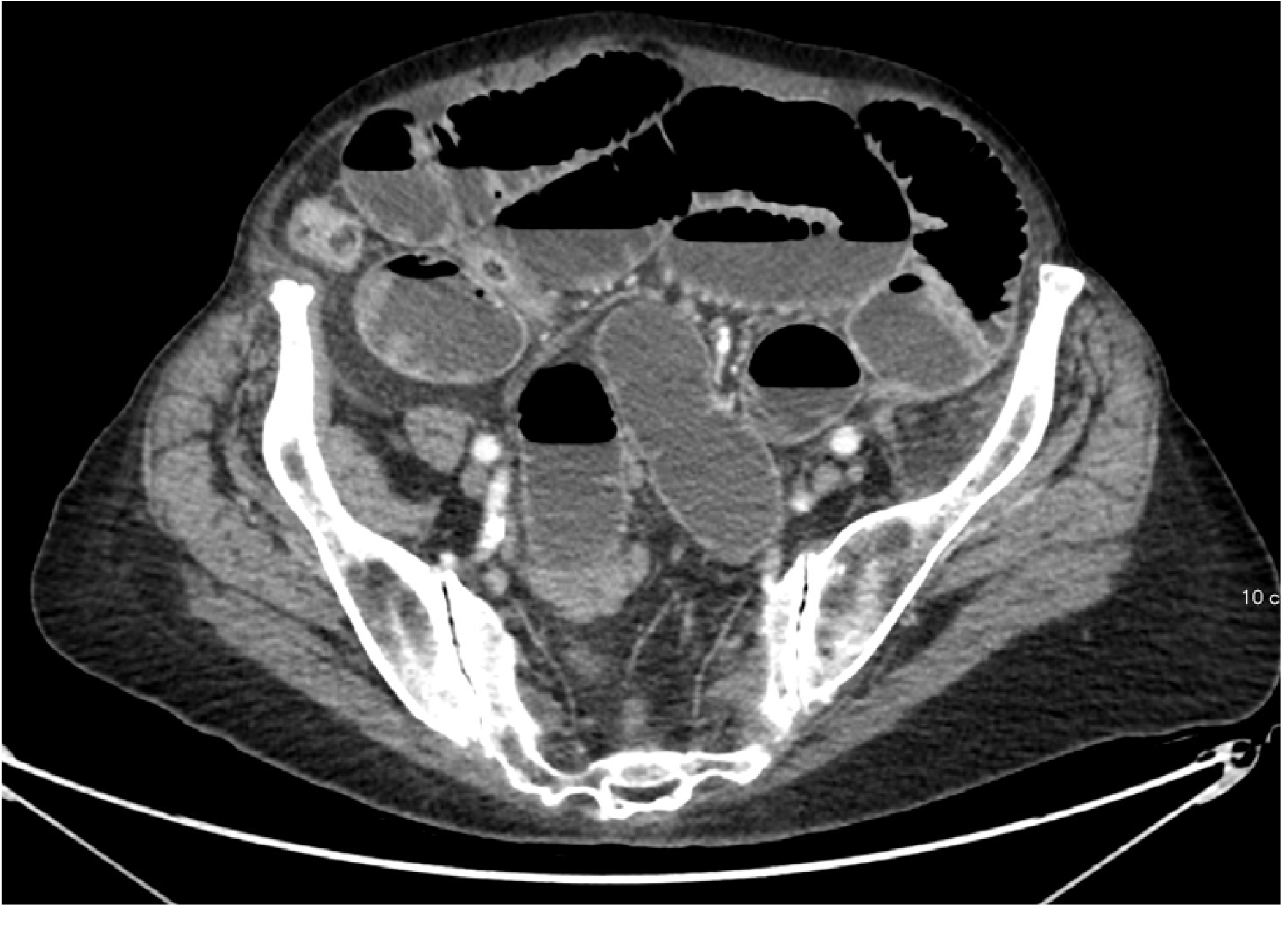
CT image of the lower abdomen with dilated small bowel loops consistent with obstruction. GPT-4V correctly identified the modality and the anatomical region but misclassified the image as normal. ‘‘‘ GPT-4V’s full answer: json } “modality”: “CT,” “anatomical_region”: “Abdomen/Pelvis,” “pathology”: “Normal” { ‘‘‘

**Figure 3.**
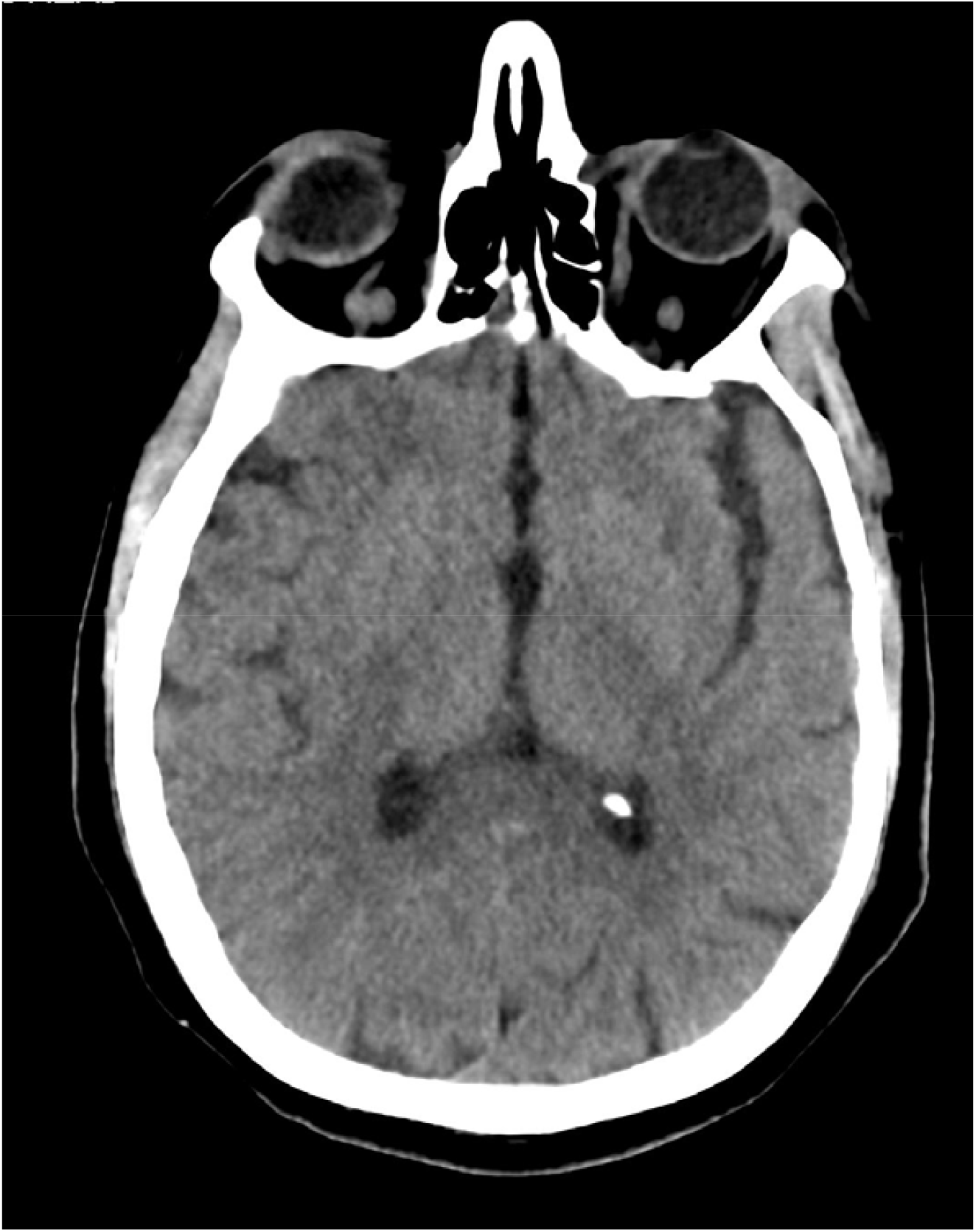
Normal CT image of the head. GPT-4V correctly identified the modality and the anatomical region but hallucinated a pathology of intracranial hemorrhage. ‘‘‘ GPT-4V’s full answer: json } “modality”: “CT,” “anatomical_region”: “head,” “pathology”: “intracranial hemorrhage” { ‘‘‘

**Figure 4.**
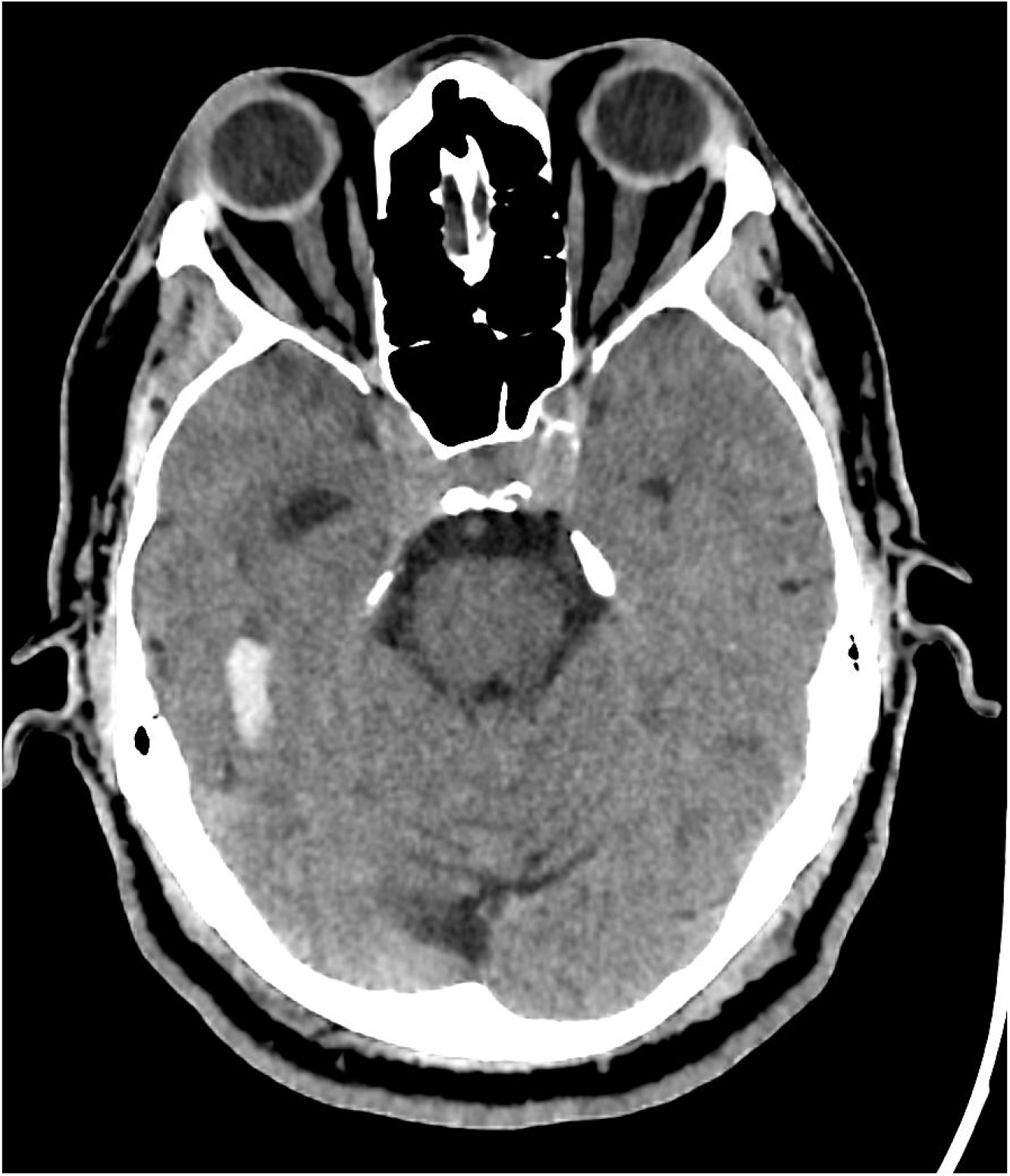
CT image of the head with a right temporal parenchymal hemorrhage. GPT-4V correctly identified the image modality, anatomical region, and pathology of “Intracranial hemorrhage”. ‘‘‘ GPT-4V’s full answer: json } “modality”: “CT,” “anatomical_region”: “Head,” “pathology”: “Intracerebral Hemorrhage” { ‘‘‘

**Figure 5.**
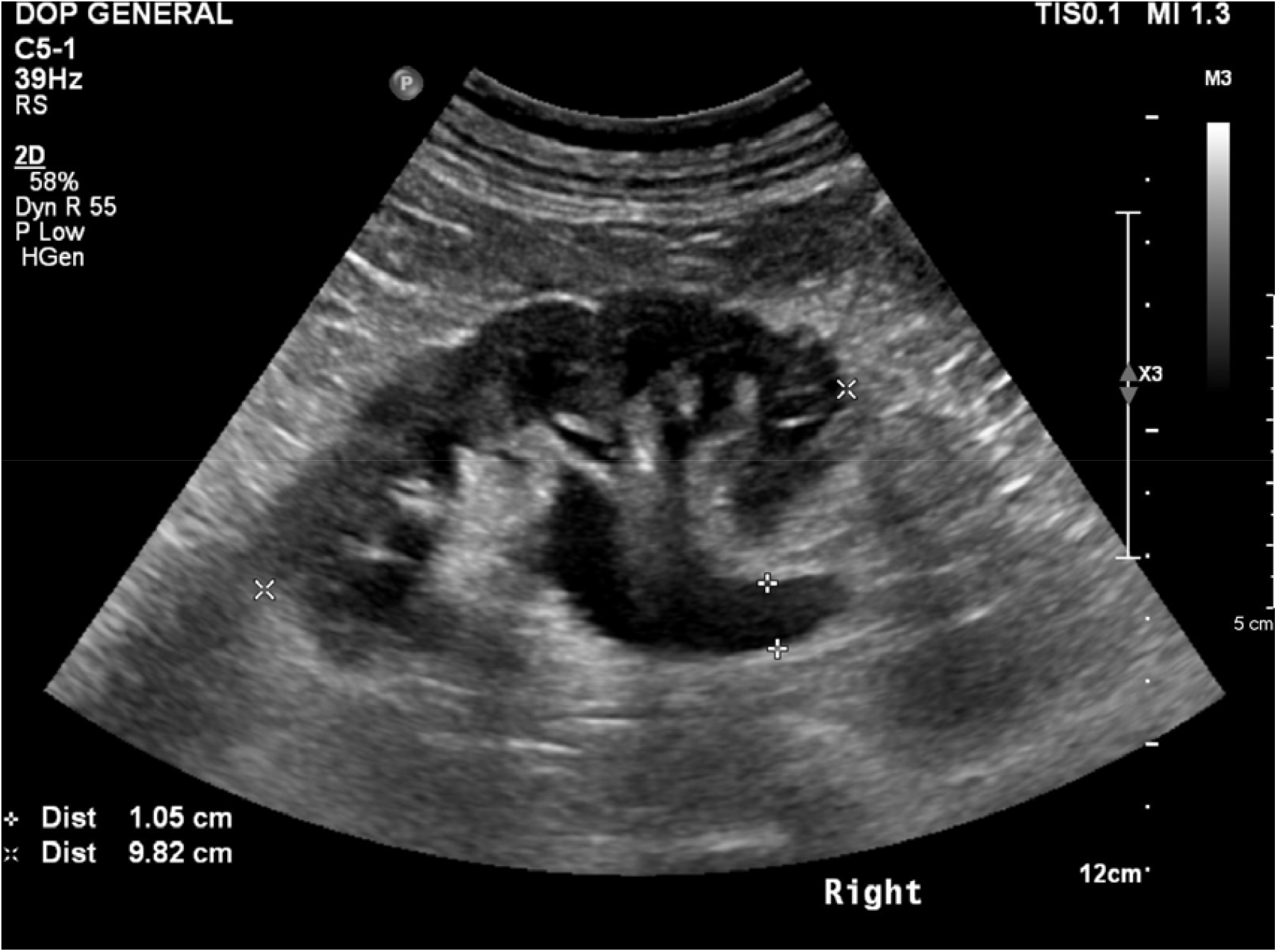
US image of the right kidney demonstrating hydronephrosis. GPT-4V identified the modality but misidentified the anatomical region as “Pelvis” and the pathology as “Cholelithiasis”. GPT-4V’s full answer: ‘‘‘json } “modality”: “Ultrasound,” “anatomical_region”: “Pelvis,” “pathology”: “Cholelithiasis” { ‘‘‘ ‘‘‘

**Figure 6.**
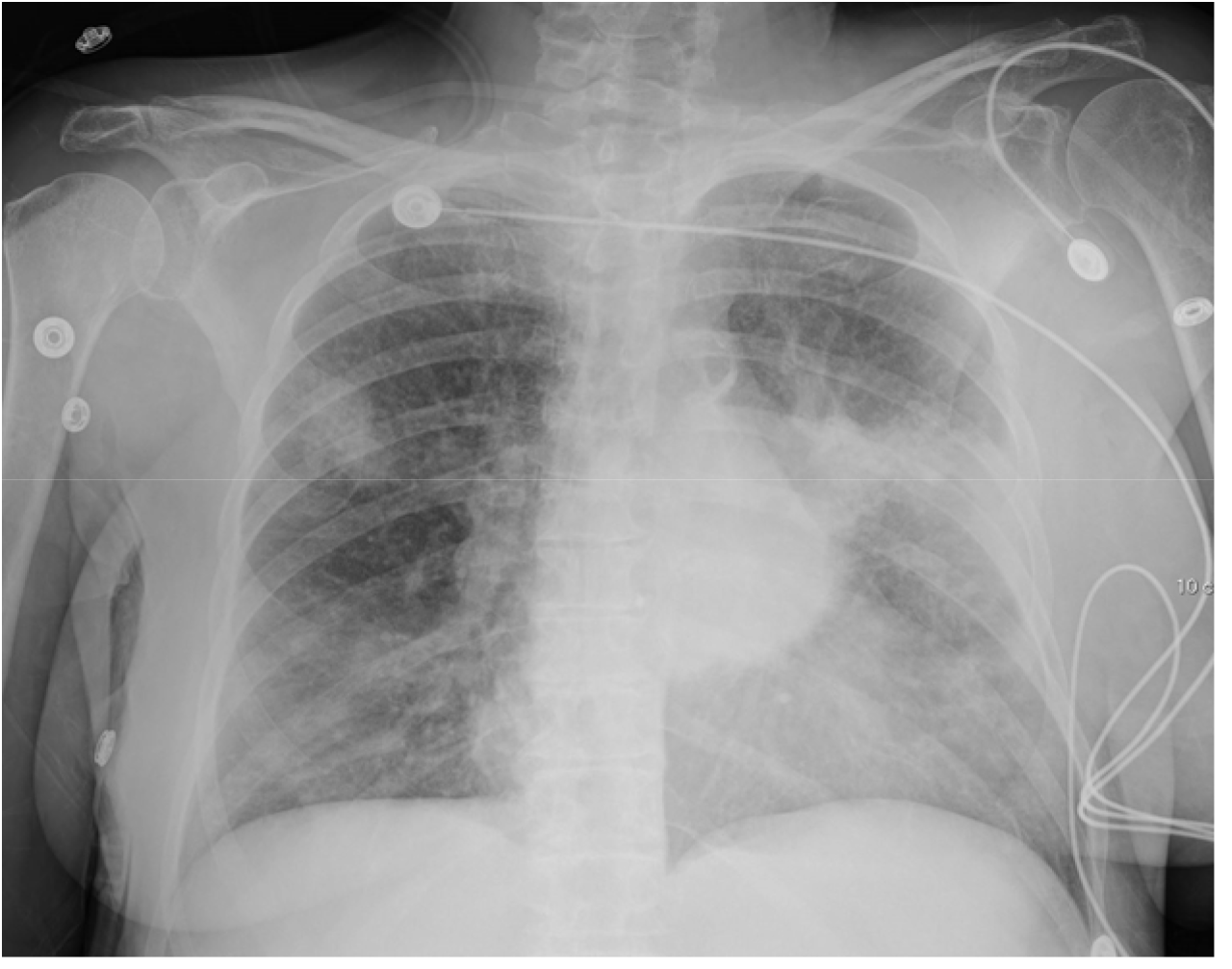
Chest x-ray with bilateral lung opacities, more prominent on the left. GPT-4V correctly identified the modality and anatomical region. Regarding the presence of a pathology, GPT-4V answered: “The presence of a heterogenous opacification in the right hemithorax with a mediastinal shift towards the left, and volume loss in the right lung, could be indicative of a large pleural effusion or a lung consolidation such as pneumonia, or a mass. Further clinical correlation and additional diagnostic imaging, such as CT, may be required for a definitive diagnosis.” When analyzing this answer, the model correctly identified the presence of opacification and a possible differential diagnosis, but with inaccuracies regarding the laterality of the pathology (right vs. bilateral) and hallucination of mediastinal shift, which is not seen in this image. GPT-4V’s full answer: json modality”: “Radiography anatomical_region”: “Chest pathology”: “The presence of a heterogenous opacification in the right hemithorax with a mediastinal shift towards the left, and volume loss in the right lung, could be indicative of a large pleural effusion or a lung consolidation such as pneumonia, or a mass. Further clinical correlation and additional diagnostic imaging, such as CT, may be required for a definitive diagnosis ‘‘‘

### Error Analysis

Error analysis across imaging modalities is detailed in **Table 4**.

**Table 4:** GPT-4 mistake types across different modalities. Identified/Total (%)

| Modality | Pathology omission and classifying case as normal | Hallucination of an incorrect pathology | Hallucinations in normal cases | Overall Hallucinations |
|---|---|---|---|---|
| **CT** | 10/53 (18.9%) | 24/53 (45.3%) | 27/46 (58.7%) | 51/99 (51.5%) |
| **X-ray** | 7/29 (24.1%) | 5/29 (17.2%) | 5/22 (22.7%) | 10/51 (19.6%) |
| **US** | 1/29 (3.4%) | 11/29 (37.9%) | 29/37 (78.4%) | 40/66 (60.6%) |
| **Total** | 18/111 (16.2%) | 40/111 (36%) | 61/105 (58.1%) | 101/216 (46.8%) |

Abbreviations: computed tomography (CT), ultrasound (US)

Hallucinations of pathologies were noted in 101/216 (46.8%) of cases. The rate of pathology hallucinations varied among modalities. The highest hallucination rate was noted in US at 40/66 (60.6%). CT scans showed an overall hallucination rate of 51/99 (51.5%). X-rays showed the lowest hallucinations rate of 10/51 (19.6%).

A recurrent error in US imaging involved the misidentification of testicular anatomy. In fact, the testicular anatomy was only identified in 1 of 15 testicular US images. Pathology diagnosis accuracy was also the lowest in US images, specifically in testicular and renal US, which demonstrated 7.7% and 4.7% accuracy, respectively.

## Discussion

This study offers a detailed evaluation of multimodal GPT-4 performance in radiological image analysis. GPT-4V correctly identified all imaging modalities. The model was inconsistent in identifying anatomical regions and pathologies, exhibiting the lowest performance in US images. The overall pathology diagnostic accuracy was only 35.2%, with a high rate of 46.8% hallucinations. Consequently, GPT-4V, as it currently stands, cannot be relied upon for radiological interpretation.

However, the moments where GPT-4V accurately identified pathologies show promise, suggesting enormous potential with further refinement. The extraordinary ability to integrate textual and visual data is novel and has vast potential applications in healthcare, and radiology in particular. Radiologists interpreting imaging examinations rely on imaging findings alongside the clinical context of each patient. It has been established that clinical information and context can improve the accuracy and quality of radiology reports[17]. Similarly, the ability of LLMs to integrate clinical correlation with visual data marks a revolutionary step. This integration not only mirrors the decision making process of physicians, but also has the potential to ultimately surpass current image analysis algorithms which are mainly based on convolutional neural networks (CNNs)[18,19].

GPT-4V represents a new technological paradigm in radiology, characterized by its ability to understand context, learn from minimal data (zero-shot or few-shot learning), reason, and provide explanatory insights. These features mark a significant advancement from the traditional AI applications in the field. Furthermore, its ability to textually describe and explain images are awe-inspiring, and with the algorithm’s improvement may eventually enhance medical education.

A preceding study assessed GPT-4V’s performance across multiple medical imaging modalities, including CT, X-ray, and MRI, utilizing a dataset comprising 56 images of varying complexity sourced from public repositories(20). In contrast, our study not only increases the sample size with a total of 230 radiological images but also broadens the scope by incorporating US images, a modality widely used in ER diagnostics.

We did not incorporate MRI due to its less frequent use in emergency diagnostics within our institution. Our methodology was tailored to the ER setting by consistently employing open-ended questions, aligning with the actual decision-making process in clinical practice. This approach reinforces and extends the findings of the previous study, corroborating their conclusion that the present iteration of GPT-4V falls short in reliability for diagnostic use and underscores the need for cautious application of AI in clinical diagnostics.

This study has several limitations. First, this was a retrospective analysis of patient cases, and the results should be interpreted accordingly. Second, there is potential for selection bias due to subjective case selection by the authors. Finally, we did not evaluate the performance of GPT-4V in image analysis when textual clinical context was provided, this was outside the scope of this study.

To conclude, despite its vast potential, multimodal GPT-4 is not yet a reliable tool for clinical radiological images interpretation. Our study provides a baseline for future improvements in multimodal LLMs and highlights the importance of continued development to achieve clinical reliability in radiology.

## Data Availability

All data produced in the present study are available upon reasonable request to the authors

## Notes

### Competing Interest Statement

The authors have declared no competing interest.

### Funding Statement

This study did not receive any funding

### Author Declarations

The IRB of Sheba Medical Center gave ethical approval for this work

### Summary of Updates

1. Increased sample size: We have substantially increased the study's sample size to 230 cases ensuring over 50 cases per modality and a balance between pathologic and normal cases. 2.Methodological elaborations and transparency: The methods section has been extensively revised to provide greater detail and clarity. This includes a more precise definition of the criteria for case selection, ensuring the focus on "clear-cut" cases, and the detailing of the exact prompts used for the AI analysis. 3.Enhanced discussion on pathology diagnostic accuracy: We have deepened the discussion regarding the overall pathology diagnostic accuracy of GPT-4V. This includes detailed methods, results, and discussion sections, along with a newly added Table 3 and Figures 1-6, which demonstrate GPT-4V's performance, including the types of errors. 4.Enhanced literature review: The discussion now includes references to updated literature.

